# An “*Infodemic*”: Leveraging High-Volume Twitter Data to Understand Public Sentiment for the COVID-19 Outbreak

**DOI:** 10.1101/2020.04.03.20052936

**Authors:** Richard J. Medford, Sameh N. Saleh, Andrew Sumarsono, Trish M. Perl, Christoph U. Lehmann

## Abstract

**Background:** Twitter has been used to track trends and disseminate health information during viral epidemics. On January 21, 2020, the CDC activated its Emergency Operations Center and the WHO released its first situation report about Coronavirus disease 2019 (COVID-19), sparking significant media attention. How Twitter content and sentiment has evolved in the early stages of any outbreak, including the COVID-19 epidemic, has not been described.

**Objective:** To quantify and understand early changes in Twitter activity, content, and sentiment about the COVID-19 epidemic.

**Design:** Observational study.

**Setting:** Twitter platform.

**Participants:** All Twitter users who created or sent a message from January 14th to 28th, 2020.

**Measurements:** We extracted tweets matching hashtags related to COVID-19 and measured frequency of keywords related to infection prevention practices, vaccination, and racial prejudice. We performed a sentiment analysis to identify emotional valence and predominant emotions. We conducted topic modeling to identify and explore discussion topics over time.

**Results:** We evaluated 126,049 tweets from 53,196 unique users. The hourly number of COVID-19-related tweets starkly increased from January 21, 2020 onward. Nearly half (49.5%) of all tweets expressed fear and nearly 30% expressed surprise. The frequency of racially charged tweets closely paralleled the number of newly diagnosed cases of COVID-19. The economic and political impact of the COVID-19 was the most commonly discussed topic, while public health risk and prevention were among the least discussed.

**Conclusion:** Tweets with negative sentiment and emotion parallel the incidence of cases for the COVID-19 outbreak. Twitter is a rich medium that can be leveraged to understand public sentiment in real-time and target public health messages based on user interest and emotion.

**Funding:** None.

## Introduction

With over 300 million monthly users, the micro-blogging platform Twitter is used increasingly to disseminate public health information and obtain real-time health data using crowdsourcing methods [1]. Researchers analyzed Twitter data to project the spread of influenza and other infectious outbreaks in real time [2]. In 2009, investigators measured the evolving interest in an Influenza A outbreak by analyzing tweet keywords and estimating real-time disease activity and disease prevention efforts [3]. During the Ebola virus (EV) outbreak in 2014, Twitter users publicized pertinent health information from media sources with peak Twitter activity within 24 hours following news events [4]. Tweet content analysis following the EV epidemic discovered that Ebola-related tweets revolved mainly around risk factors, prevention, disease trends, and compassion [5]. Similarly, the 2015 Middle Eastern Respiratory Syndrome (MERS) outbreak, disease spread was found to be correlated with Twitter activity, promoting Twitter as a potential surveillance tool for emerging infectious diseases [6]. During the Zika virus epidemic, Twitter was used to study significant changes in travel behavior due to mounting public concerns [7].

Recognizing Twitter’s potential to inform and educate the public, governmental agencies such as the World Health Organization (WHO) and the Centers for Disease Control and Prevention (CDC) have adopted the use of Twitter and other social media. In the first 12 weeks of the Zika outbreak in late 2015, the WHO Twitter account was retweeted over 20,000 times, demonstrating its widespread impact on disseminating health information [8].

In December 2019, the first diagnosis of a novel, emerging coronavirus, formally named severe acute respiratory syndrome coronavirus (SARS-CoV-2), was made in Wuhan City, Hubei Province, China. In subsequent weeks, the coronavirus’s rapid spread garnered increasing media coverage and public attention. Press coverage further heightened on January 21, 2020 when the CDC activated its Emergency Operations Center and the WHO began publishing daily situation reports. Subsequent travel limitations and cancellations, large-scale quarantine of Chinese residents, and numerous international index cases generated significant interest by the general public [9]. However, there is limited insight into the main topics discussed and the sentiment of the general public over time.

We postulate that analysis of the content and sentiments expressed on Twitter in the early stages of the coronavirus disease 2019 (COVID-19) pandemic will parallel the spread of the disease and can aid understanding of the effect of the outbreak on the sentiments, beliefs, and thoughts of the general public. Such understanding would enable large-scale opportunities for education and appropriate information dissemination about public health recommendations.

## Methods

### Overview

We constructed a list of hashtags related to COVID-19 to search for relevant tweets during a two-week interval from January 14^th^ to 28^th^, 2020. We extracted the tweets using Twitter’s advanced search application programing interface (API) and stored them as plain text. We identified themes and analyzed the frequency of associated keywords including infection prevention practices, vaccination, and racial prejudice. We performed a sentiment analysis using the text of tweets to identify each tweet’s emotional valence (positive, negative, or neutral) [10] and predominant emotion (anger, disgust, fear, joy, sadness, or surprise) [11]. Finally, we performed topic modeling using an unsupervised machine learning method to identify and analyze related topics over time within the corpus of the tweets [12].

### Data Collection

From January 14^th^ to 28^th^ 2020, a random sample of tweets in the English language was extracted using Twitter’s API and its advanced search tool (https://twitter.com/search-advanced). The Twitter stream was filtered in accordance with Twitter’s advanced search algorithm resulting in a representative subset of tweets. The dates were chosen to include one week of data before and after the activation of the Emergency Operations Center by the Centers for Disease Control and Prevention [13] and the release of the first WHO situation report [14]. Hashtags used for identification of 2019-nCoV related tweets included *#2019nCoV, #coronavirus, #nCoV2019, #wuhancoronavirus*, and *#wuhanvirus (COVID-19* and *SARS-COV-2* were not coined until Feb 19, 2020). Collected metadata from tweets included nineteen variables, of which eight were used in our analysis: tweet text, time stamp, if the tweet had a reply, if the tweet was a reply, if the tweet was unique or a retweet, if the tweet included an image, if the tweet included a link, the number of tweet likes, number of retweets, and number of replies.

### Data processing, transformation, and exploration

We performed all data processing and analysis using Python software, version 3.6.1 (Python Software Foundation) and RStudio version 1.2.1335 (R Foundation for Statistical Computing). We compared the COVID-19-related tweets per hour with the number of newly confirmed cases over each 24-hour period and completed descriptive statistics for the collected metadata. To analyze tweets, we extracted the plain text from the original message and stripped out web addresses, Twitter hyperlinks, and punctuation. For all but the sentiment analysis, we removed stop words (words commonly found in a document of little analysis value e.g., “for”, “the”, “is”), converted text to lowercase, and lemmatized words (changing different forms of a word to its root form e.g., “viruses” to “virus” or “went” to “go”). We transformed the words in tweets into a vector of individual words and two-word phrases (i.e., unigrams and bigrams respectively). We removed terms present in less than five tweets and two words that were found in greater than ten percent of tweets (“case” and “people”) decreasing the dictionary from 626,614 terms to 38,823 terms.

Using a word cloud, we visualized the top three hundred words with larger font size representing greater frequency. We used a subset of keywords to identify tweets related to three common infection prevention and control (IPC) strategies, vaccination, and racial prejudice. Appendix Section A details the keywords used. We analyzed the incidence of these tweets over time and manually reviewed a random 10% subset to validate content, evaluate narratives present, and explore examples of misinformation.

### Sentiment Analysis

Emotional valence describes emotions that refer to the intrinsic attractiveness or aversiveness of a subject like events, objects, or situations [15]. We analyzed the emotional valence of tweets separately using four commonly used methods through the *Syuzhet* R package [10]. Because each method uses a different scale, we normalized scores to detect the polarity of tweets as positive, negative, or neutral. For the emotion analysis, we used recurrent neural networks to label a primary emotion for a document according to Ekman’s emotional classification (i.e., anger, disgust, fear, joy, sadness, or surprise) [11]. We trended the findings by visualizing the daily number of tweets labeled with each emotional valence and each emotion over the two-week period and comparing their rate of change by tweets per day.

### Topic Modeling

A Latent Dirichlet Allocation (LDA) [12] model (gensim Python package [16]) automatically generates topics from observations (in our case, from tweets) and groups similar observations to one or more of these topics using the distribution of words. We iteratively trained multiple LDA models using different numbers of topics to maximize a topic coherence score (which measures the degree of semantic similarity between high scoring words in the topic). Selecting the highest coherence score resulted in the use of the LDA model with ten topics. Adhering to convention, we presented the top fifteen terms (a common number of terms used in analyzing topics in LDA models) that contributed to each topic group and manually labeled a theme for each topic (Figure 6a). We then visualized the topic model using a t-distributed Stochastic Neighbor Embedding (t-SNE) graph [17], which embeds high-dimensional data (i.e., ten dimensions given ten topics) into a graphable two-dimensional space where similar tweets are grouped together (Figure 6b). We created an interactive visualization of the t-SNE to qualitatively evaluate the change in topics over time.

## Results

### Tweet Frequency

A total of 126,049 tweets (of which, 123,407 were unique) from 53,196 unique users were collected during the study period. The most prevalent identification hashtag found was *#coronavirus* followed by *#wuhancoronavirus* present in 82% and 13% of tweets, respectively. The tweets accumulated 114,635 replies, 1,248,118 retweets, and 1,680,253 likes. In the first week of our analysis, the number of COVID-19-related tweets remained stable with less than 100 tweets per hour. The number of tweets per hour began increasing on January 20th, and reached as many as 250 per hour by January 21st and continued to grow with a peak of over 1,700 tweets per hour by January 28th, 2020. This trend closely tracked the number of newly confirmed COVID-19 cases (Figure 1).

**Figure 1:**
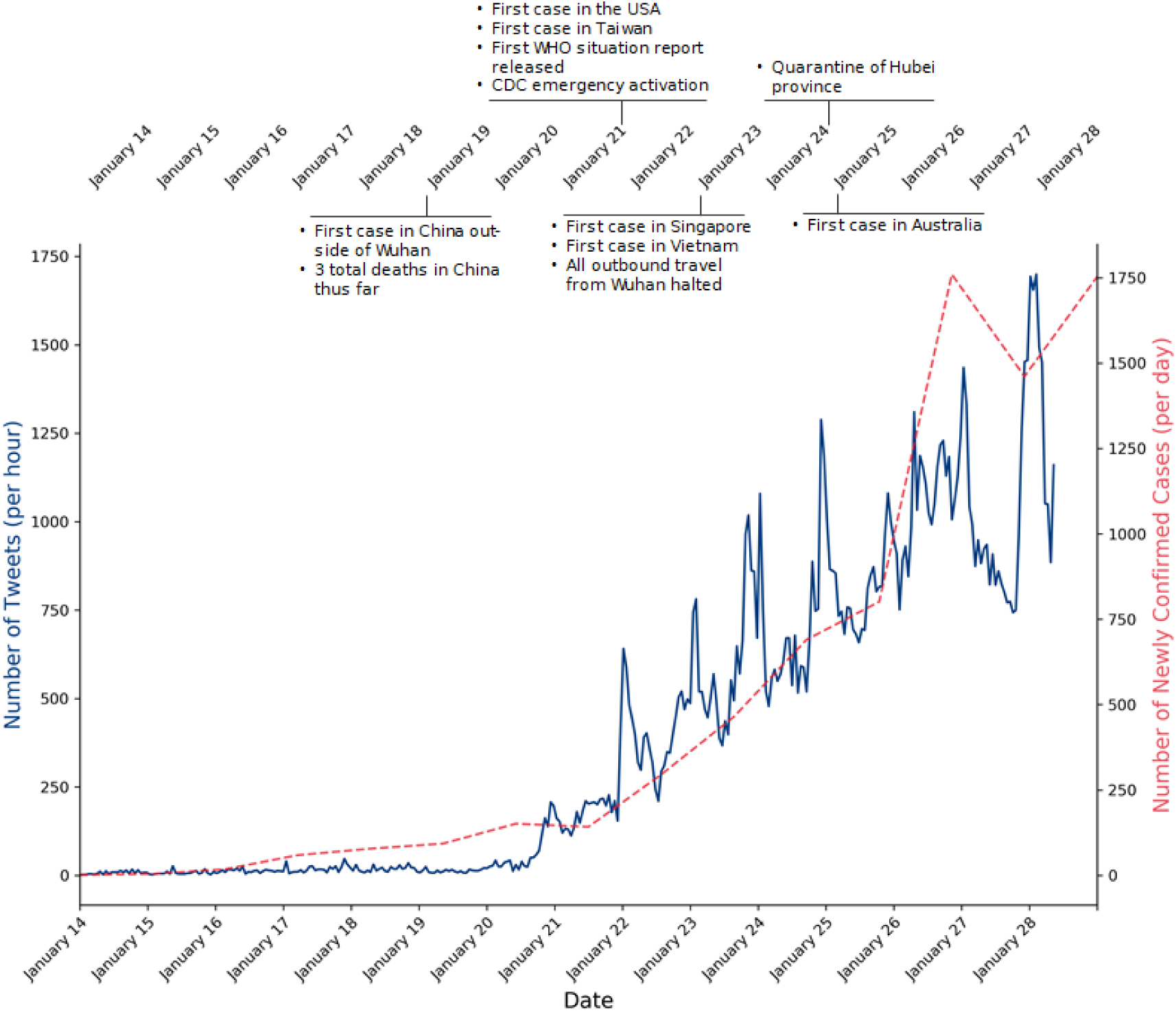
Number of COVID-19-related tweets (left y-axis) and number of newly confirmed coronavirus cases (right y-axis) over time.

### Common Expressions

Collected tweets contained 2,877,816 words and 15,955,720 characters. The most common word in our analysis was ‘outbreak’, numbering 11,549 times (Figure 2). The other top fifteen most commonly used words and their frequency in descending order were: ‘spread’ (11,290), ‘health’ (9,734), ‘confirm’ (6,897), ‘death’ (5,819), ‘city’ (5,662), ‘report’ (5,662), ‘first’ (5,431), ‘world’ (5,244), ‘travel’ (5,049), ‘hospital’ (4,405), ‘infect’ (4,388), ‘SARS’ (4,133), ‘mask’ (3,996), ‘patient’ (3,981), and ‘country’ (3,885).

**Figure 2:**
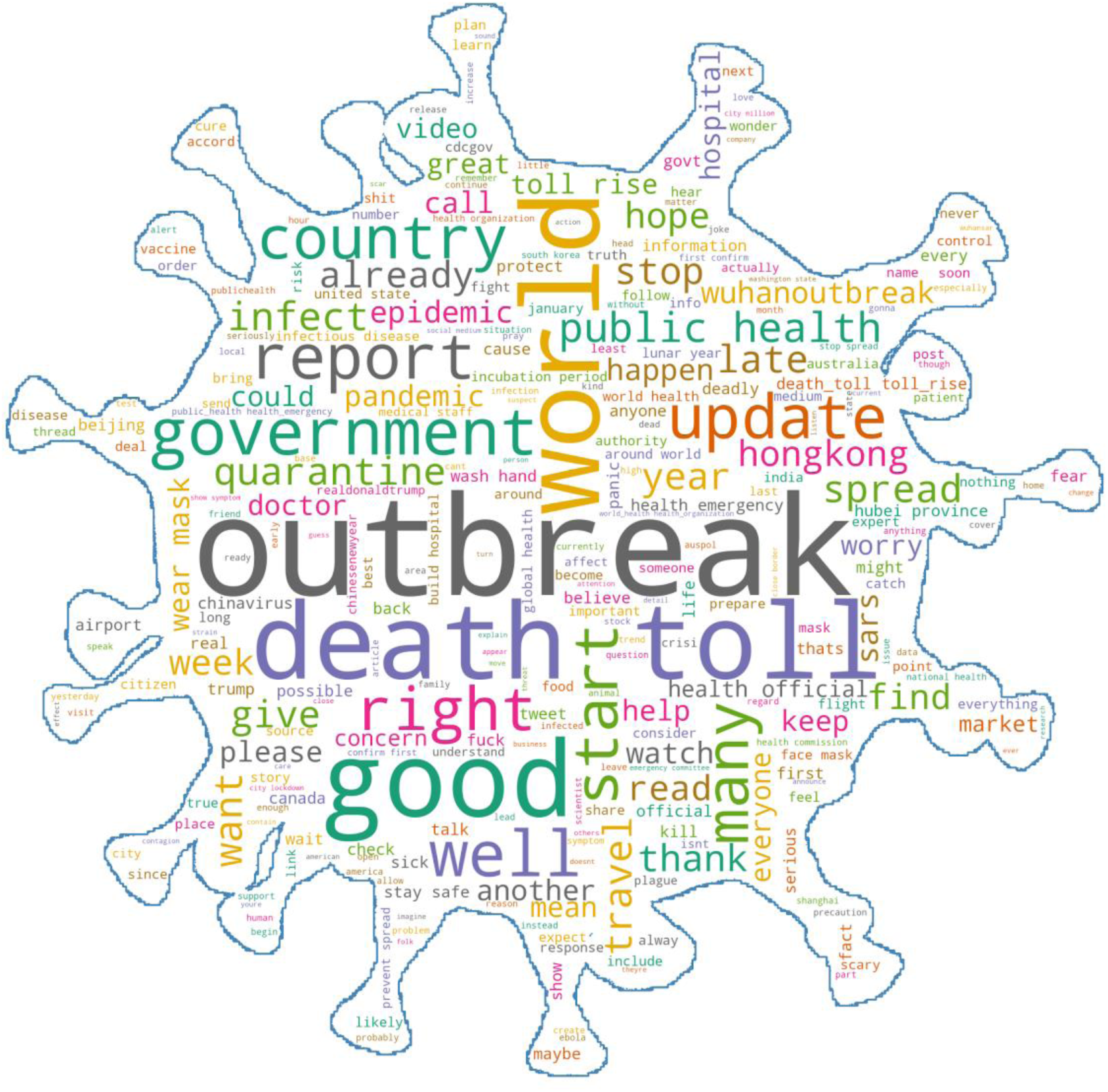
Word cloud showing the top 300 words used in tweets related to COVID-19.

### Infection Prevention and Control

Prior to January 20th, our analysis showed very few tweets related to infection prevention and control (IPC) followed by a steady increase starting January 21st (Figure 3). Isolation-related tweets were the most prevalent followed by mask and hand hygiene. Coinciding with the quarantine of the Hubei province, isolation-related tweets disproportionately increased on January 24. All IPC subgroups increased over time but their ranking did not change. IPC-related content was present in 4.8% of tweets. Discussions of prevention techniques, shortage of protective gear, dissemination of health information, and large-scale quarantine were most common. Tweets with reference to vaccinations were found in 1.2% of total tweets and increased at a slower rate than IPC-related tweets overall. The most prevalent vaccine-related tweets were about vaccine availability, vaccine development, and advocacy to receive the influenza vaccine.

**Figure 3:**
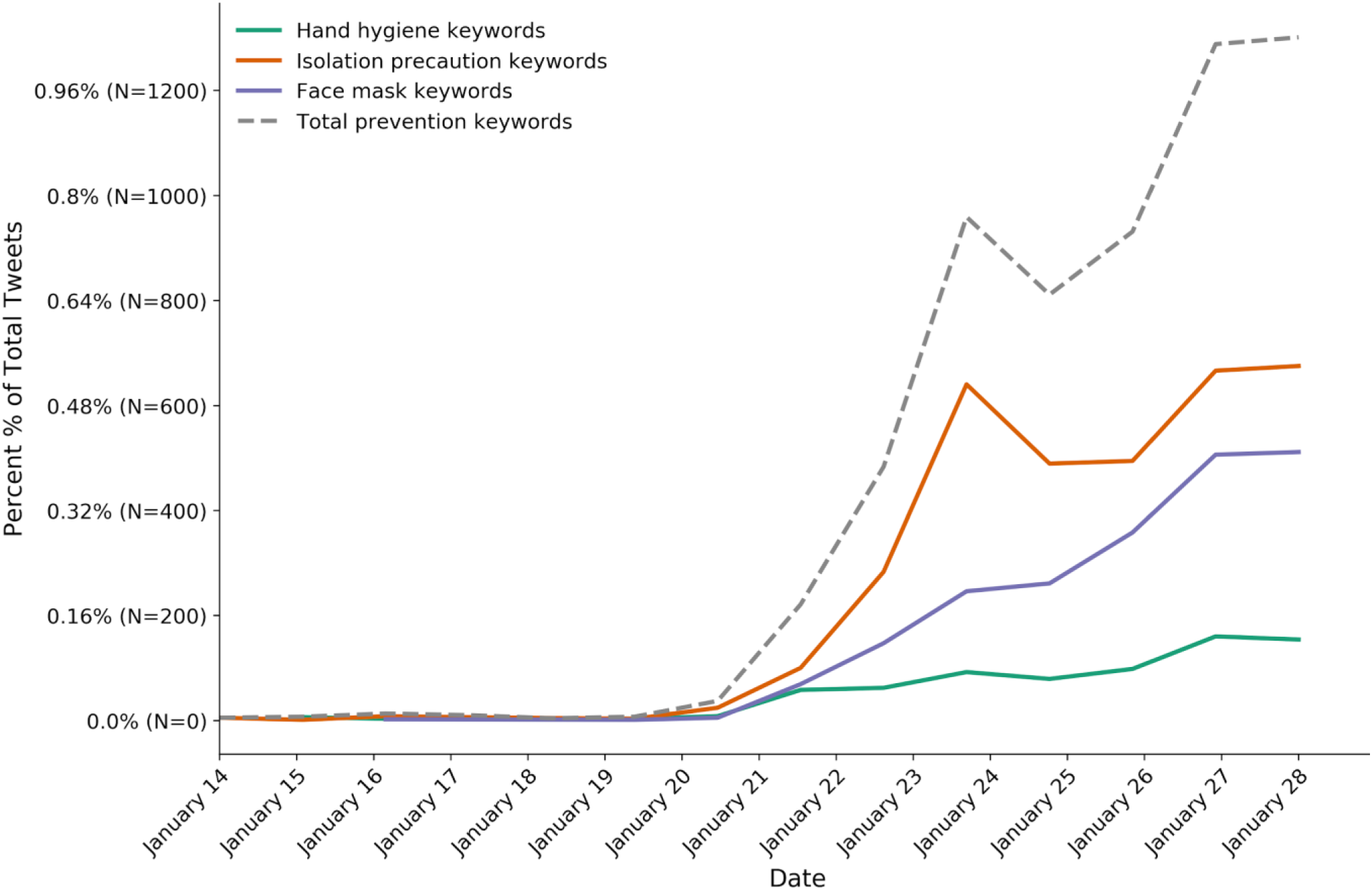
Daily number of tweets related to infection protection and its sub groups of isolation/quarantine, masks, and hand hygiene.

### Emotions

Fear was the most common emotion expressed in nearly 49.5% of all tweets with topics ranging from fear of infection, death, and inability to travel as well as emotional distress and fear regarding the effect on the economy and politics. [Examples: “*Coronavirus: Virus fears trigger Shanghai face mask shortage*” and “*Oil falls below $60 as China coronavirus fears accelerate*”] Surprise was the second most common emotion present in 29.3% of tweets. [Example: “*The Wuhan virus is more critical than expected! Don’t forget to wear [a] face mask(surgical mask)!*”]. Anger followed and included themes of inadequate governmental reactions, isolation and quarantine, lack of supplies, and lack of information. [Examples: “*Wuhan coronavirus: Hong Kong police, protesters clash as anger erupts over proposal to use housing block as quarantine site*” and “*11 million city on a lockdown!!!*”]. The least common predominant emotions found in tweets were sadness, joy, and disgust (Figure 4a). We analyzed tweets for positive, neutral, or negative emotional valence. Tweets with a negative emotion were more common than neutral and positive tweets and increased at a faster rate over time (Figure 4b). More sample tweets are included in Appendix Figure 1.

**Figure 4:**
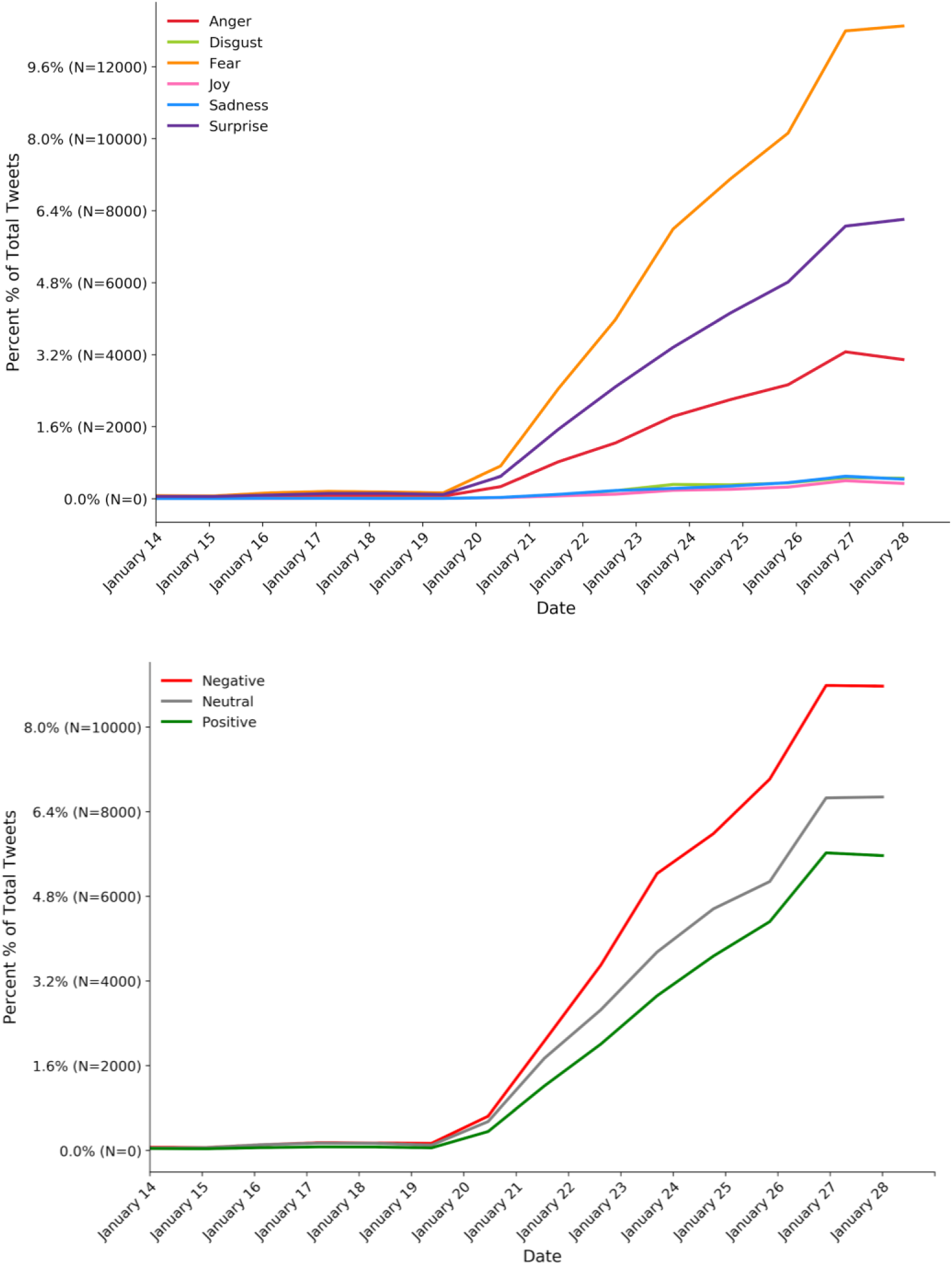
Analysis of a) tweet emotions (fear, anger, surprise, sadness, joy, and disgust) and b) emotional valence over time.

### Racial Prejudice

Tweets related to xenophobic content or racial prejudice were largely absent at the start of our observation period. Subsequently, their number increased daily and paralleled the number of new COVID-19 cases (Figure 5). Racial prejudice was present in 0.54% of tweets. The offensive nature of these tweets prohibits examples, but data are available upon request to the corresponding author.

**Figure 5:**
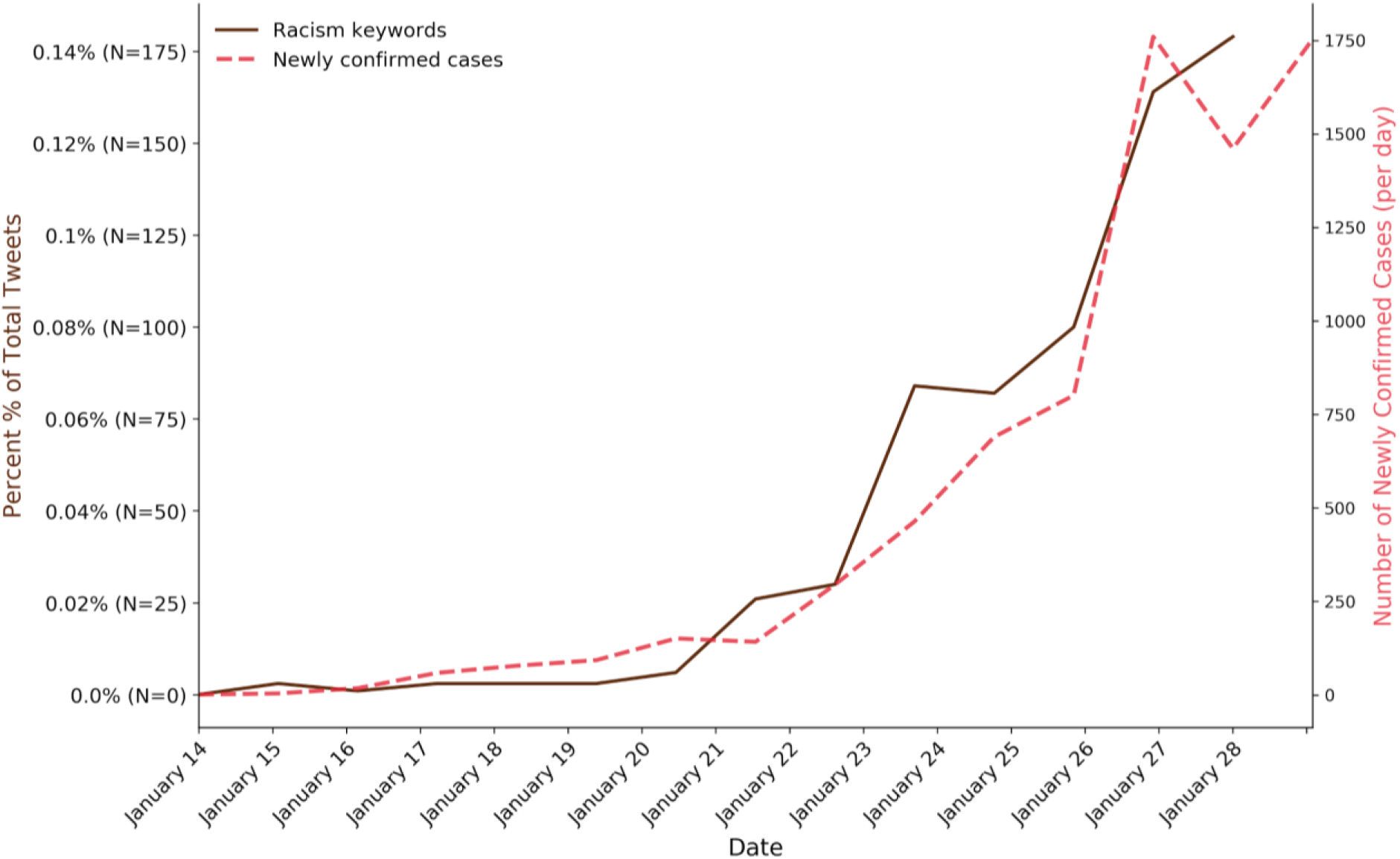
Analysis of keywords related to racial prejudice discussion over time compared to the number of cases over time.

### Topic Modeling

Topic modeling identified ten themes that are recorded in Figure 6a. Keywords are listed in order of weight in forming the abstract topics found within the text. A tweet may include multiple topics, but typically has one predominant topic. The most common predominant topic was the economic and political impact, followed by government response to the virus, then discussion of the outbreak and its development and transmission. The least common topics included index cases, the public health response, and healthcare provision. Other topics included the number of cases and death as well as prevention and large-scale quarantine. An interactive visualization of tweet themes showing their development by day is available at https://ssaleh2.github.io/Early_2019nCoV_Twitter_Analysis/; hovering over a node will show the tweet text and the day it was posted (please note the figure is slow to load and the slider on top allows navigation through time). Figure 6b shows three screenshots from the visualization.

**Figure 6a.**
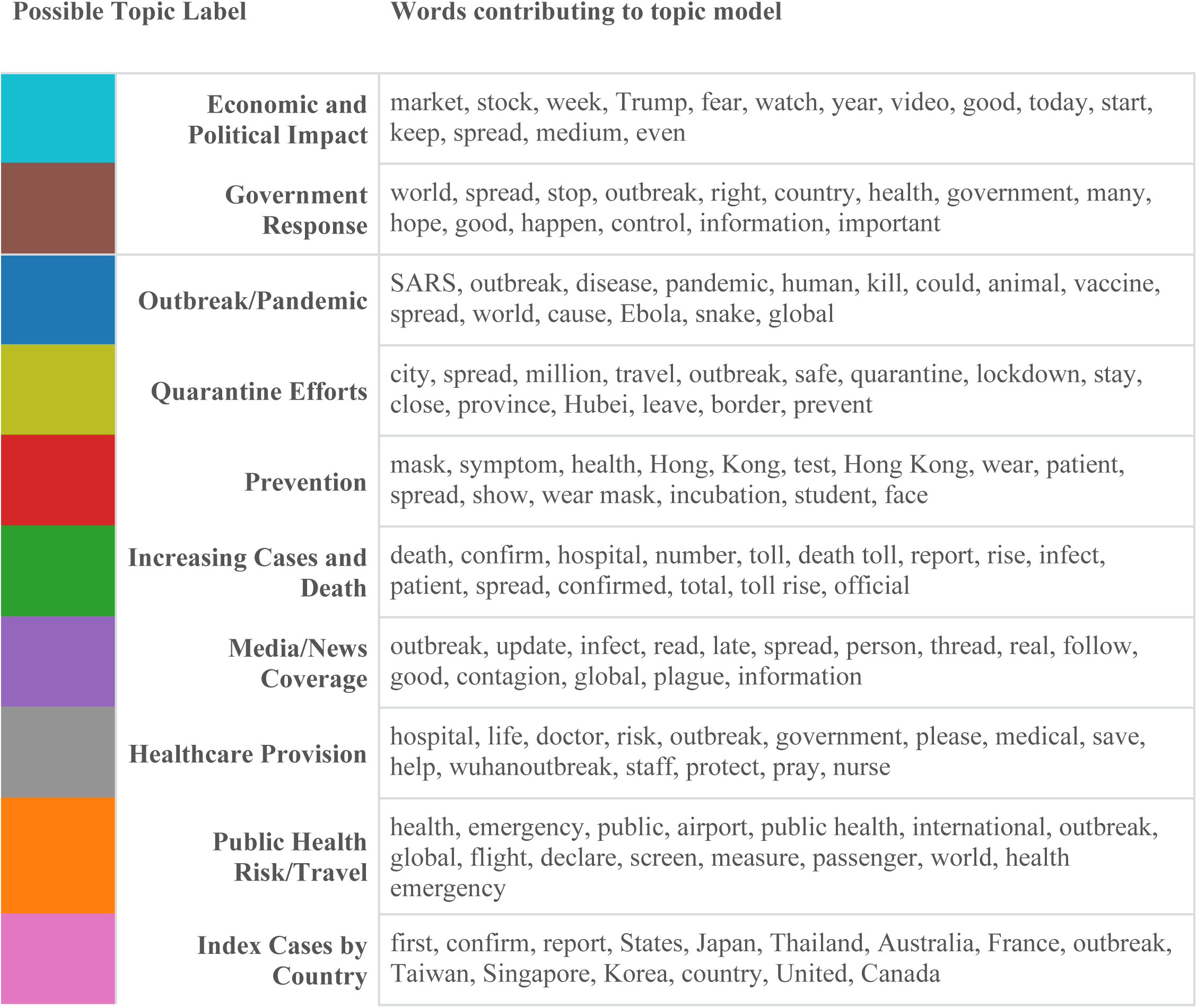
The fifteen terms (in order of weighting) that contributed to each abstract topic with their potential theme labels. The topics are ordered by frequency. Colors for each topic correspond to those in Figure 6b. Topic labels were assigned by the authors.

**Figure 6b.**
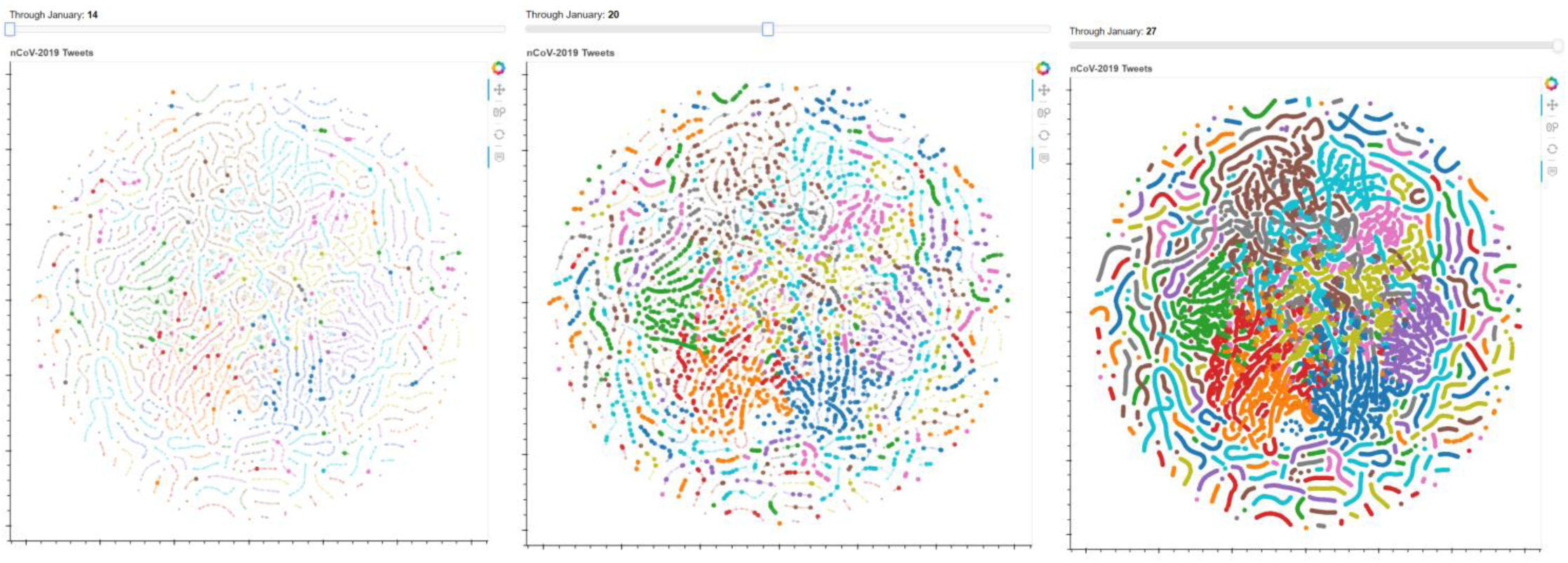
A t-distributed Stochastic Neighbor Embedding (t-SNE) graph (which embeds high-dimensional data into a two-dimensional space where similar tweets are grouped together) that visualizes the topics in Figure 6a as labeled by color and how they change over time. The full interactive visualization is available at https://ssaleh2.github.io/Early_2019nCoV_Twitter_Analysis/; please note the visualization is slow to load. Each node represents an individual tweet and only tweets posted through the day highlighted on the slider are shown in the foreground, while all tweets in the study period are shown in the background. Hovering over a node will show the tweet text and the day it was posted. Depicted here are three screenshots for January 14 (day 0), January 20 (day 6), and January 27 (day 13).

Major themes clustered in the center while more obscure tweets displayed in the periphery. As tweets may include multiple topics, there is visible cross-over between topic clusters in the visualization. Topic clusters that included themes of outbreak and its transmission, public health risk, and index cases were discussed from the start of the study period, while discussion of quarantine effects, economic and political impact, and government response increased significantly in the second week of the study period.

## Discussion

In this study, we demonstrate significant persistent increases in overall Twitter activity, tweets with negative sentiment and emotions, and racially charged content for the COVID-19 outbreak from January 21, 2020 onward. We show that the frequency of tweets was associated with the number of infected individuals for the early stages of the COVID-19 outbreak. Tweets predominantly showed negative sentiment and were linked to emotions of fear primarily, as well as surprise and anger. While tweets with misinformation and societal prejudice were present, tweets were also significantly used to disseminate valuable public health information. These data may help medical experts and public health officials to identify types of communication and messaging that may allay emotion and decrease misinformation.

Emotions have been shown to alter how we think, decide, and solve problems especially in highly charged situations of outbreaks [18]. Further, “[p]atients’ perception […] of our health care system […], informs, and is, their reality” [19]. For public health officials, governments, and health care industry leaders, understanding public sentiment and reaction to infectious outbreaks is crucial to predict utilization of healthcare resources and compliance with public health and infection prevention measures. Twitter allows us access to the thoughts and emotions of millions of users and permits efficient and real-time analysis of these sentiments on important health care topics like the ongoing COVID-19 outbreak.

Surveillance programs for emerging and highly dangerous infections are difficult and labor intensive [20]. Leveraging the information of the crowd by analyzing social media posts offers a simple and in the case of the COVID-19 outbreak, a surprisingly realistic view of the extent of the public health emergency. Despite collecting only tweets in English, the number of daily tweets paralleled the number of newly diagnosed cases even though most of these early cases were in China.

Global infectious outbreaks such as the COVID-19 pandemic may become the culture medium for a societal illness: social segregation and racial prejudice. As previously reported in other outbreaks, such as EV [21] and the Severe Acute Respiratory Syndrome (SARS) [22], there is significant discrimination surrounding acquisition of these viruses toward individuals based on ethnicity and race. Similarly, we found a substantial number of tweets related to racial prejudice in our study sample. The increase of these tweets mirrored the rise in newly confirmed COVID-19 cases. Their overall rate was relatively low at 0.54%, but taken at scale, this is concerning as the press has reported racist or discriminating behavior towards Chinese-American and other Asian-American citizens [23,24].

Twitter is currently the most popular social media platform for healthcare communication. Skepticism of its utility has been long discussed, with Twitter’s opponents often citing misinformation and the inability to process high volumes of information [25]. We found evidence of misinformation and hyperbole in tweets and reported online (Examples: “*People are literally dying on the streets of China [*…]”, “*The new fad disease ‘coronavirus’ is sweeping headlines. Funny enough, there was a patent for the coronavirus was (sic) filed in 2015 and granted in 2018*”, and “*Tesla Models S and X hospital grade HEPA filters may help prevent coronavirus infection*”). [More sample tweets are available in Appendix Figure 1.] Social media companies such as Facebook, Google, and Twitter have taken on the responsibility of acting as stewards of information related to COVID-19 by removing false information and redirecting web traffic to reputable websites [26]. The account of the user, who tweeted the misleading patent information above was subsequently suspended [27]. Twitter Singapore adjusted their search prompt to show links to authoritative health sources like the WHO and Ministry of Health for the COVID-19 outbreak [28]. As evident in our analysis, misinformation is prevalent on Twitter, but it represented a small fraction of the communication.

Further, it is important to point out that scientists and government officials also contributed to the dissemination of false information during this outbreak. A description of transmission in a prominent journal falsely reported that an asymptomatic person infected four others with coronavirus [29]. Researchers failed to interview the index case, who later reported that she had been symptomatic [30]. A since withdrawn scientific article falsely claimed that COVID-19 has four pieces of sequence in its genetic code not found in other coronaviruses and speculated that the virus could be genetically engineered [31].

Overall, the majority of tweets were intended to disseminate knowledge. Crowdsourcing has been shown to be an enormously powerful and expedient way of achieving educational tasks [32]. The desire of the crowd to use a tool like Twitter to obtain and disseminate information offers the opportunity to change the narrative and educate millions of people. Since the outbreak started, the WHO has educated the public with a steady stream of tweets [33]. Some tweets analyzed were related to infection prevention measures (hand washing, coughing into your sleeve, self-isolation), but these were still the minority, present in less than 5% of tweets.

From a public health perspective, the ability to analyze Twitter feeds in real-time and the potential to individually target segments of the population with high-impact messages based on their information needs and sentiment could be an extremely powerful tool, potentially more effective than any other communication medium. To date, bots (autonomous programs able to interact with computer systems or users) have often been used on Twitter for advertising or to promulgate malicious or false content [34,35]. However, public health and governmental organizations like the WHO or the CDC should invest in this new technology. For example, deploying autonomous tools that identify tweets by users who are fearful of contracting COVID-19 could be used to send individually targeted messages that provide reassurance and education on preventive measures such as handwashing. Tailoring automatic responses to the sentiments and content of tweets has the potential to engage more Twitter users on public health topics and to redirect the discussion to useful, accurate information.

### Limitations

This study had several limitations. First, we used a non-comprehensive list of hashtags that was limited by knowledge of trending hashtags and the imagination of the authors. We may have missed alternative terminology or misspellings and may have introduced some selection bias in the tweets we analyzed. For example, *#wuhanoutbreak* was not included, but arose as a weighted term in our topic modeling. Conversely, *#coronavirus* may have identified tweets related to other infections such as Severe Acute Respiratory Syndrome. Second, despite the large number of tweets analyzed (>126K), we collected and analyzed only a subset (1%) of all tweets, which may also introduce some selection bias. However, using the Twitter API, we were assured that the sample constituted a representative subset of the entire stream. Third, we targeted tweets in the English language; thus, our conclusions may not be generalizable to other countries where English is not the predominant language. Lastly, we recognize that ascribing topic themes based on a subset of weighted terms has opportunity for labeling bias. To mitigate that, two authors designed the topic model and a separate set of authors labeled the topic themes.

### Conclusions

We show that the frequency of tweets was associated with the number of infected individuals for the early stages of the COVID-19 pandemic. Tweets predominantly showed negative sentiment and were linked to emotions of fear primarily, as well as surprise and anger. While tweets with misinformation and societal prejudice were present, tweets were also significantly used to disseminate valuable public health information. Twitter offers novel opportunities to public health and governmental agencies to not only track public perception of infectious outbreaks, but also to target messages of a public health nature based on user interest and emotion.

## Data Availability

All data available upon request. An abridged dataset with the tweets included are available at the link below.

https://github.com/ssaleh2/Early_2019nCoV_Twitter_Analysis

## Availability of data and materials

The data that support the findings of this study are available upon request.

## Authors’ contributions

Study concept and design: RM, SS, CL, AS; Data acquisition and extraction: SS; Data analysis: RM, SS; Interpretation of data: RM, SS, CL; Manuscript preparation: all authors. All authors read and approved the final manuscript.

